# Advanced and more advanced revascularization in STEMI patients: which is better?

**DOI:** 10.1101/2024.03.11.24304135

**Authors:** Yahui Li, Chunxia Zhao, Xindi Yue, Ling Zhou, DaoWen Wang, Feng Wang

## Abstract

**Objective:** To compare the long-term prognosis of patients who experienced acute ST-segment elevation myocardial infarction and underwent either late percutaneous coronary intervention (PCI) within a period of 2 days to less than 1 week or more advanced PCI within 1 week to less than 1 month.

**Methods:** We enrolled 198 patients from Tongji Hospital of Tongji Medical College, Huazhong University of Science and Technology, between June 2019 and August 2022. These patients had experienced acute ST-segment elevation myocardial infarction and underwent either late PCI or more advanced PCI. Long-term follow-up was conducted through outpatient clinic visits or telephone interviews. The study endpoints included all-cause death, nonfatal myocardial infarction, and New York Heart Association class IV heart failure. We utilized the Kaplan-Meier method to illustrate the cumulative incidence of endpoint events in both patient groups. Statistical significance in survival differences was assessed using the log-rank test. Additionally, the Cox proportional risk model was employed to analyze whether the timing of late revascularization procedures had an impact on the long-term prognosis of the patients.

**Results:** Among the 198 patients included in the study, 108 underwent late PCI, while 90 underwent more advanced PCI. The majority were male (73.74%), with an average age of 62 ± 13 years. The follow-up period averaged 20 (15, 28) months, and all patients successfully completed the follow-up process. Analysis based on the Kaplan-Meier method revealed that the incidence of all-cause death [11.1% vs. 5.6%, P=0.165], nonfatal myocardial infarction [7.4% vs. 7.8%, P=0.922], New York Heart Association class IV heart failure [2.8% vs. 3.3%, P>0.999], and the composite endpoint [18.5% vs. 14.4%, P=0.444] were not statistically significant between the late PCI and more advanced PCI groups. After adjusting for factors like left ventricular ejection fraction, renin-angiotensin system inhibitors, β-blockers, and statins, the results still indicated no statistically significant differences between the two groups in terms of rates for all-cause death, recurrent myocardial infarction, New York Heart Association class IV heart failure, and composite endpoints (P=0.05).

**Conclusion:** This study’s 20 (15, 28) months follow-up suggests that patients experiencing acute ST-segment elevation myocardial infarction have a comparable prognosis regardless of whether they underwent late or more advanced PCI.

---

Acute ST-segment elevation myocardial infarction (STEMI) stands as a frequent and critical cardiovascular emergency, often leading to significant mortality and disability among individuals with coronary heart disease^1^. Prompt and effective re-opening of the arteries affected by the infarction is a crucial life-saving intervention that significantly enhances the prognosis of STEMI patients. Percutaneous coronary intervention (PCI) emerges as the favored reperfusion strategy^2^. As outlined in the 2021 ACC/AHA/SCAI guidelines for coronary revascularization, emergency PCI is designated as a Class I recommendation for STEMI onset within <12 hours, while it’s classified as a Class IIa recommendation for occurrences between 12 to 24 hours. In essence, the recommended window for emergency PCI is within 24 hours of symptom onset^3^. However, due to various medical constraints and other factors, numerous patients with acute myocardial infarction in China are unable to undergo timely PCI. In real-world scenarios, a small subset of STEMI patients still received PCI 48 hours after the disease onset. There remains a scarcity of studies that compare the timing of late-stage revascularization procedures in STEMI patients. Thus, the primary objective of this study is to assess and compare the long-term prognosis of patients with STEMI who underwent PCI at later and more advanced stages.

## Information and methods

1. Subjects: This retrospective cohort study included patients diagnosed with ST-segment elevation myocardial infarction (STEMI) who underwent late or more advanced percutaneous coronary intervention (PCI) at Tongji Hospital, Tongji Medical College, Huazhong University of Science and Technology, between June 2019 and August 2022. Diagnostic criteria for STEMI^4^: (1) Persistent anterior precordial dullness lasting more than 20 minutes, unrelieved by nitroglycerin or rapid-acting heart pills. (2) ST-segment elevation in 2 or more relevant leads: ≥ 0.2mV in thoracic leads or ≥ 0.1mV in limb leads, or new complete left or right bundle-branch block. Inclusion criteria: Patients diagnosed with STEMI who underwent PCI 48 hours or more after symptom onset. Exclusion criteria: (1) Severe heart valve disease. (2) Acute pulmonary edema or severe congestive heart failure [New York Heart Association (NYHA) cardiac function class 3 or 4 or cardiogenic shock]. (3) Patients with STEMI undergoing emergent PCI or coronary artery bypass graft (CABG). (4) Patients with non-ST-segment elevation myocardial infarction undergoing emergent PCI or CABG. (5) Patients with other comorbidities significantly impacting life expectancy, such as neoplasms, severe hepatic, renal, pulmonary, endocrine, neurologic, or hematologic disorders.

Our study adhered to the principles of the Declaration of Helsinki. Prior to participation, each individual provided signed informed consent. The study procedures were approved by the Medical Ethics Committee of Tongji Hospital, Tongji Medical College, Huazhong University of Science and Technology(TJ-IRB202401016).

2. Surgical procedures and perioperative medication were managed collaboratively by the cardiologist and the patient. The cardiologist thoroughly discussed the potential risks associated with surgery and considered the patient’s preferences and input from their family while determining the optimal timing for the procedure. Medication protocols were tailored based on the cardiologist’s expertise and the patient’s individual condition. Unless medically contraindicated, all patients received an optimized medication regimen, including aspirin/indobufen, clopidogrel/ticagrelor, anticoagulants if necessary, renin-angiotensin system inhibitors, beta-blockers, and lipid-lowering therapy.

3. Collection of clinical data involved extracting information from patients’ medical records and hospitalization test results. This included: Patient Demographics and Past Medical History: Gender, age, and medical history encompassing hypertension, diabetes mellitus, hyperlipidemia, and chronic renal insufficiency. Test and Examination Data: BMI, temperature, heart rate, systolic and diastolic blood pressure, white blood cell count, red blood cell count, hemoglobin, neutrophils, lymphocytes, platelet count, alanine aminotransferase, aspartate aminotransferase, lactate dehydrogenase, creatinine, predicted glomerular filtration rate, total cholesterol, triglycerides, high-density lipoprotein (HDL) cholesterol, low-density lipoprotein (LDL) cholesterol, blood potassium, blood glucose, hs-CRP, blood sedimentation rate, CTnI, NT-proBNP, and left ventricular ejection fraction(LVEF). Medication and Treatment Details: Oxygen therapy, noninvasive and invasive respiration, aortic balloon counterpulsation, extracorporeal membrane pulmonary oxygenation, temporary pacemaker, and a range of medications such as aspirin, indobufen, clopidogrel, ticagrelor, statin, ezetimibe, PCSK9i, β-blocker, Angiotensin-Converting Enzyme Inhibitors/Angiotensin receptor blocker/Angiotensin receptor neprilysin inhibitors(ACEI/ARB/ARNI), nitrate, calcitonin antagonist, ivabradine, trimetazidine, coenzyme Q10, diuretics, digitalis, and low-molecular-weight heparin. Intraoperative Information: Offending vessel, infarct localization, presence of combined total occlusive lesions, intraoperative slow flow/no reflow, thrombus aspiration, and intraoperative medications like tirofiban, nicorandil, and nitroglycerin. Outcome Measures: Time to vessel opening, total hospital costs, duration of hospital stay, and disease regression.

4. Follow-up and study endpoints: Patients underwent regular follow-ups post-surgery conducted at the cardiology clinic, ward visits, or via telephone consultations scheduled at intervals of 1, 3, and 6 months, as well as 1, 2, and 3 years. If patients exhibited post-surgical symptoms with objective evidence of ischemia, they were admitted for coronary angiography. The study endpoints encompassed all-cause mortality, nonfatal myocardial infarction, and New York Heart Association (NYHA) class IV heart failure.

5. Statistical methods: Statistical analysis was performed using SPSS 22.0 software and R 4.3.1 software. Continuous variables following a normal distribution were presented as mean ± standard deviation (̄x ± s), and intergroup comparisons were conducted using the independent sample Student t-test. For continuous variables not conforming to a normal distribution, data were described as median with the interquartile range (M (Q1, Q3)), and between-group comparisons were conducted using the Mann-Whitney U test. Categorical variables were presented as cases with percentages (%), and comparisons between groups were assessed using the chi-square test. Survival analysis was described using the Kaplan-Meier method, and differences in survival were evaluated using the log-rank test to ascertain statistical significance. The Cox proportional hazard model was utilized to calculate the risk ratio (HR) and its 95% confidence interval (CI).

Furthermore, multifactorial adjustment was implemented to analyze the impact of various factors on patients’ prognosis, discerning whether there existed a disparity in the effect of delayed PCI versus more advanced PCI on prognosis. Statistical significance was determined at a P value of <0.05. In instances where data were missing less than <30%, K-nearest neighbor interpolation was employed.

## Results

1. Comparison of baseline data between the two groups (Table 1): This study included a total of 198 consecutive patients diagnosed with STEMI who underwent PCI either in late or more advanced stages. Among these, 108 cases were categorized in the late PCI group (Group A), whereas 90 cases belonged to the more advanced PCI group (Group B). The cohort comprised 146 (73.74%) male patients with an average age of (62±13) years. Comparative analysis revealed statistically significant differences in WBC, Neut, PLT, TG, and ESR between the two groups (all P < 0.05); however, these variations did not hold clinical significance. Specifically, the advanced PCI group exhibited higher levels of AST and C-TnI, an increased necessity for oxygen therapy, and a higher likelihood of requiring intraoperative thrombus aspiration and tirofiban usage. Additionally, the median time to disease onset before PCI was 3 (2, 5) days in the late PCI group and 11 (8, 17) days in the more advanced PCI group, indicated as median and interquartile range.

**Table 1:**
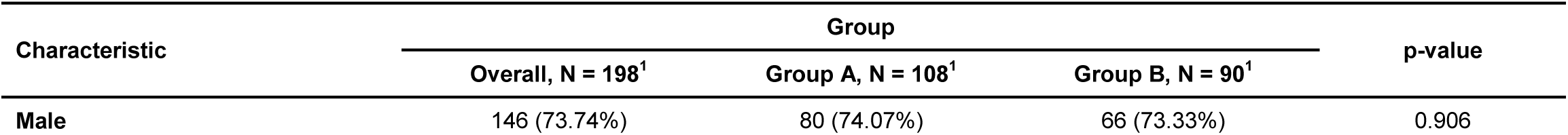
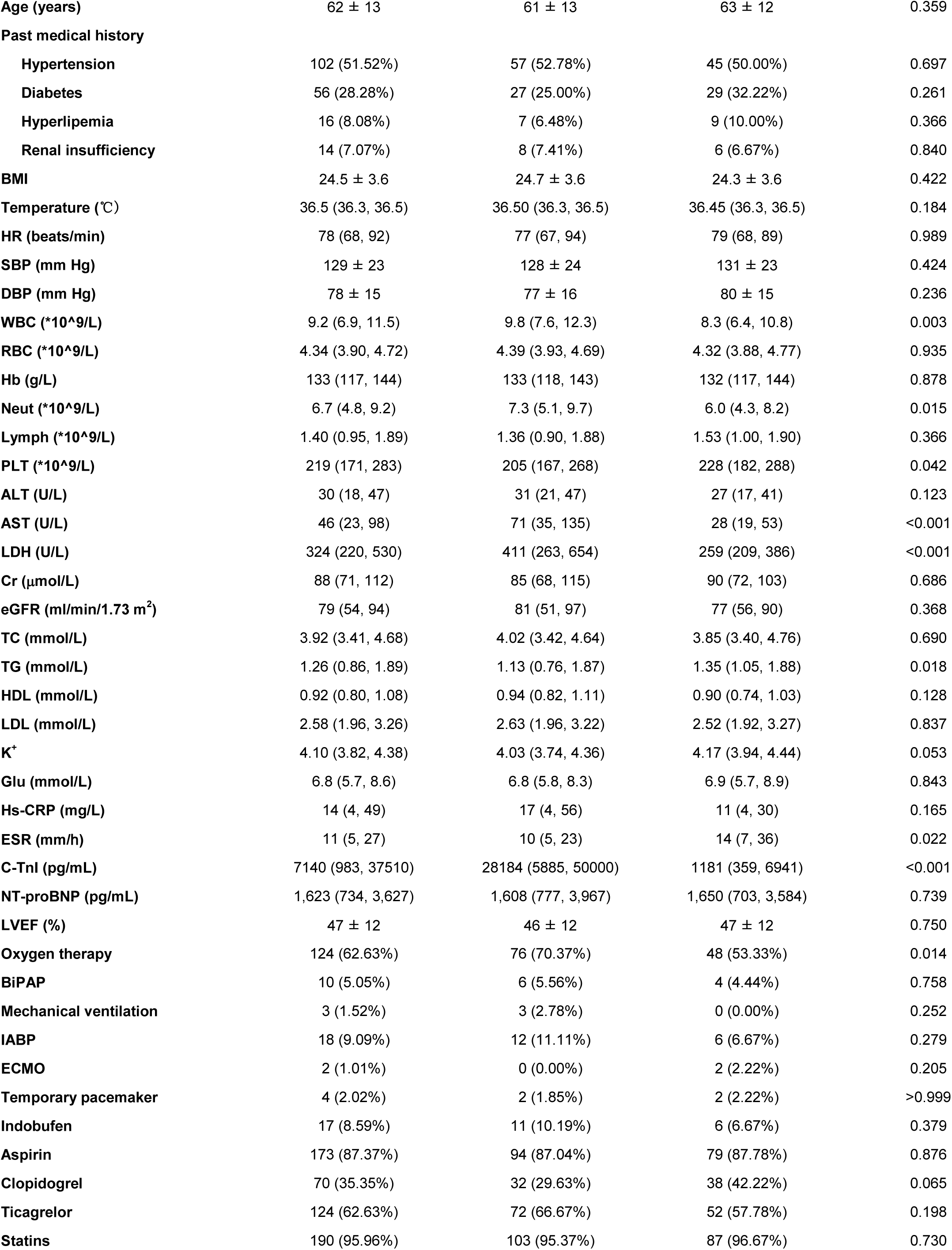
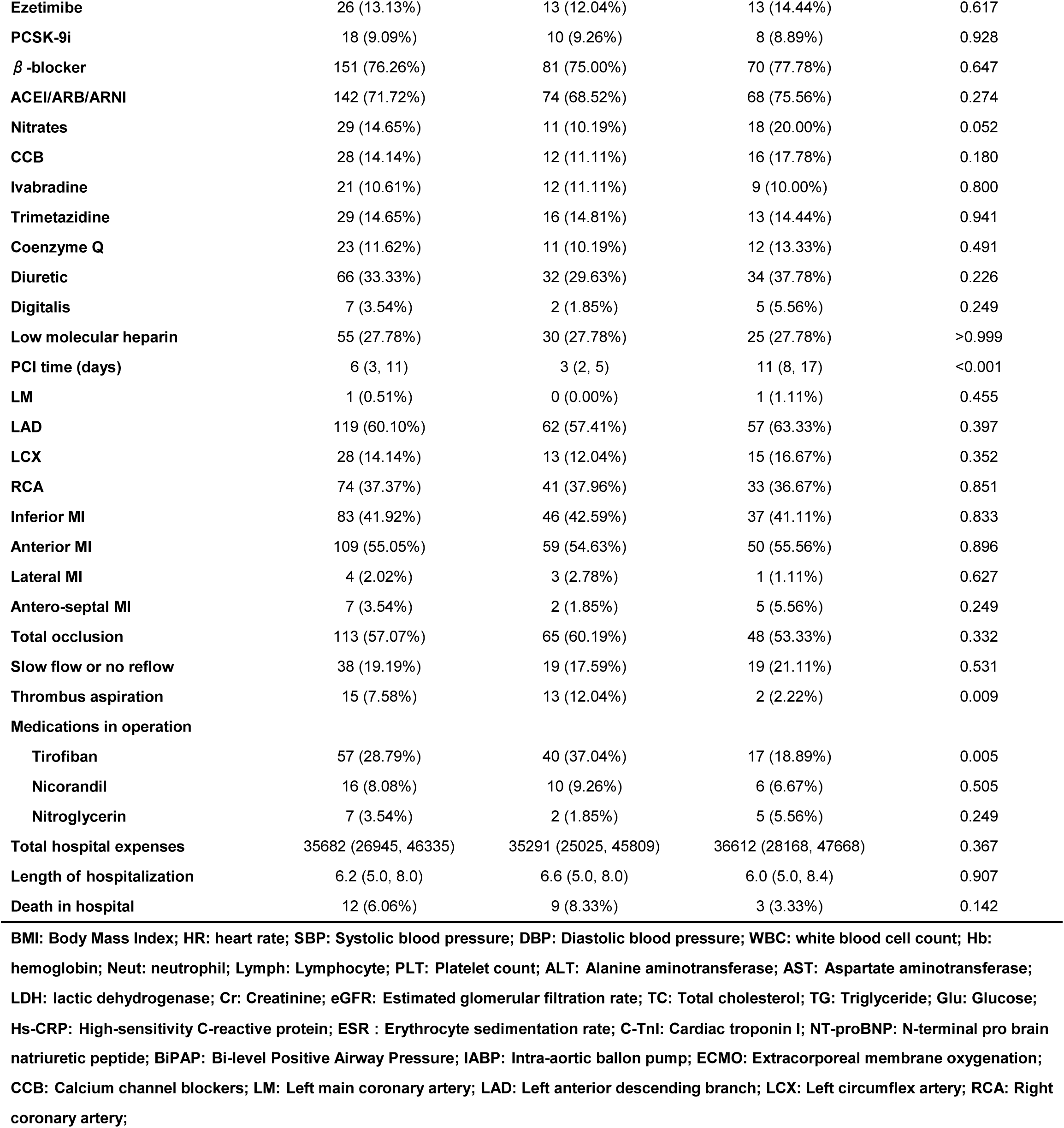
Patient demographics and baseline characteristics.

2. Comparison of prognosis between the two groups: Throughout the 20-month follow-up period (with a median duration of 20 months and interquartile range of 15 to 28 months), all patients successfully completed their scheduled follow-ups. During the follow-up period, 17 patients died (8.6% of the total cohort), comprising 12 individuals (11.1%) from the late PCI group and 5 individuals (5.6%) from the more advanced PCI group. Regarding recurrent myocardial infarctions, a total of 15 cases (7.5%) were reported, with 8 instances (7.4%) in the late PCI group and 7 occurrences (7.8%) in the more advanced PCI group. Moreover, 6 patients experienced cardiac class IV heart failure, comprising 3 cases each from both the late PCI group (2.8%) and the more advanced PCI group (3.3%).

The disparities in all-cause death, nonfatal myocardial infarction, NYHA class IV heart failure, and composite endpoints between the two groups did not display statistical significance (all P>0.05, as detailed in Table 2 and Figure 1). Subsequent correction for the four influencing factors—LVEF, ACEI/ARB/ARNI medication, β-blocker use, and Statins—indicated comparable outcomes between the two groups. Specifically, the late PCI and more advanced PCI groups exhibited comparable rates for all-cause death (HR=0.69, 95% CI 0.23-2.09, P=0.51), recurrent myocardial infarction (HR=1.14, 95% CI 0.41-3.17, P=0.81), and incidence of cardiac class IV heart failure (HR=1.49, 95% CI 0.24-9.18, P=0.67). Similarly, the composite endpoint analysis showed no statistically significant differences between the two groups (HR=0.94, 95% CI 0.45-1.94, P=0.86).

**Figure 1.**
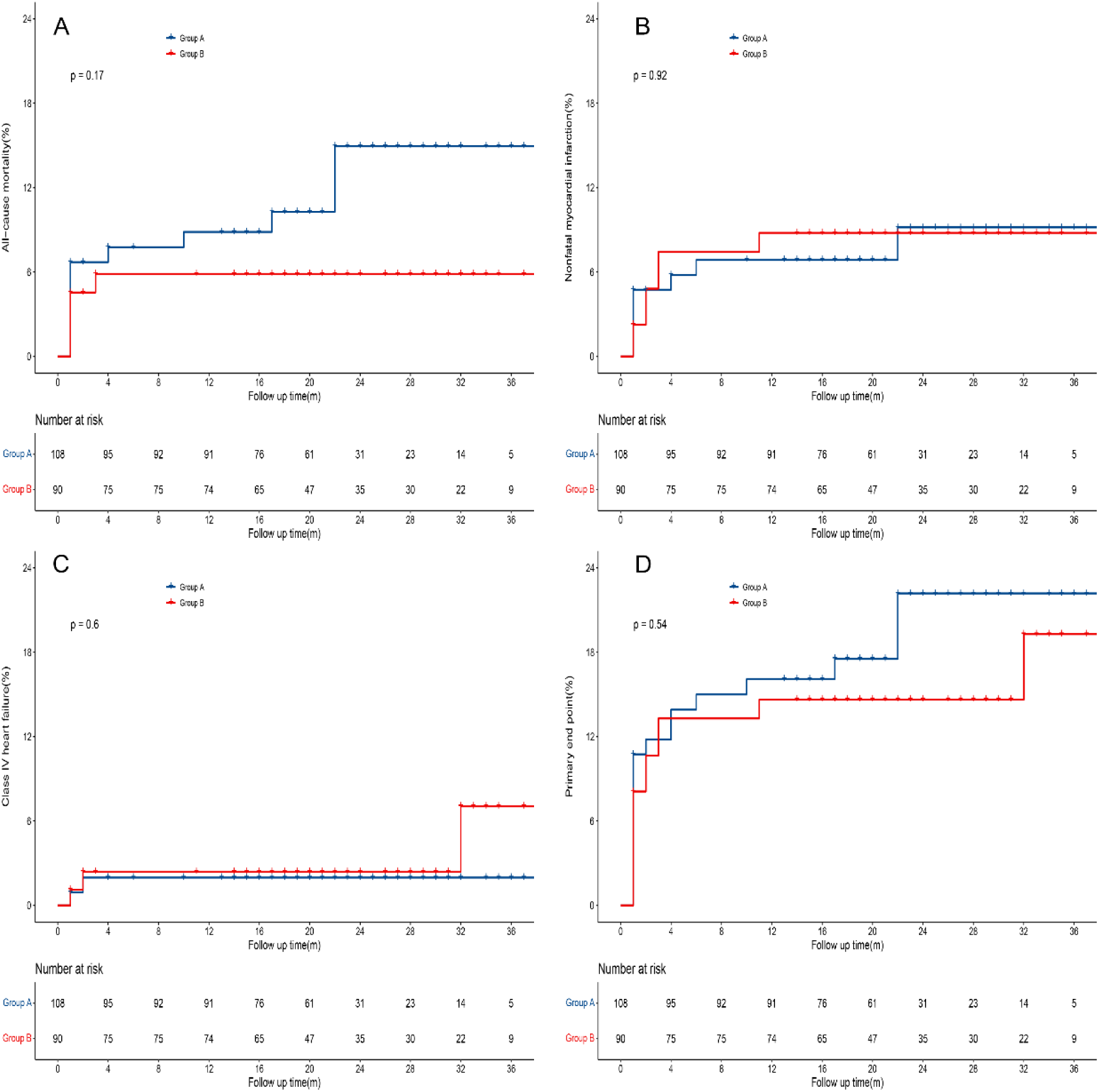
Kaplan-Meier survival curves for STEMI patients treated with different surgical timing. In Panel A, the estimated cumulative event rates for death from all causes in the Group A and Group B, respectively, were 8.5% and 5.7% at 12 months, 13.9% and 5.7% at 24 months, and 13.9% and 5.7% at 36 months. In Panel B, the estimated cumulative event rates for nonfatal reinfarction in the two groups, respectively, were 6.6% and 8.4% at 12 months, 8.8% and 8.4% at 24 months, and 8.8% and 8.4% at 36 months. In Panel C, the estimated cumulative event rates for NYHA class IV heart failure in the two groups, respectively, were 2.0% and 2.4% at 12 months, 2.0% and 2.4% at 24 months, and 2.0% and 6.8% at 36 months. In Panel D, the estimated cumulative event rates for primary end point in the two groups, respectively, were 14.9% and 13.6% at 12 months, 19.9% and 13.6% at 24 months, and 19.9% and 17.5% at 36 months. The P values for the estimated cumulative event curves at 36 months were calculated with the use of the log-rank test.

**Table 2.** Comparison of clinical events in patients with STEMI treated with different surgical timing [cases (%)]

| Endpoint indicator | Group | Group | P value | HR<sup>a</sup>(95%CI) | P value<sup>a</sup> |
| --- | --- | --- | --- | --- | --- |
|  | A(n=108) | B(n=90) |  |  |  |
| All-cause death | 12(11.1%) | 5 (5.6%) | 0.165 | 0.69 (0.23-2.09) | 0.51 |
| Nonfatal myocardial infarction | 8 (7.4%) | 7 (7.8%) | 0.922 | 1.14 (0.41-3.17) | 0.81 |
| NYHA class IV heart failure | 3 (2.8%) | 3 (3.3%) | >0.999 | 1.49 (0.24,9.18) | 0.67 |
| Composite endpoint | 20 (18.5%) | 13 (14.4%) | 0.444 | 0.94 (0.45,1.94) | 0.86 |

<sup>a</sup>:adjusted LVEF, ACEI/ARB/ARN, $\beta$ -blocke, and Statins

## Discussion

The 2018 European Society of Cardiology guidelines for myocardial revascularization advocate considering a routine direct PCI strategy for patients presenting late (between 12 to 48 hours) with symptoms, with a level IIa recommendation and level B evidence^5^. However, the guidelines advise against performing PCI in asymptomatic, stable STEMI patients more than 24 hours after symptom onset if there is complete occlusion of the infarcted artery and no indication of severe ischemia^3^. These recommendations stem from research primarily driven by the OAT trial^6^and the DECOP trial^7^. The OAT trial’s findings revealed that compared to optimal pharmacologic therapy, PCI did not decrease the incidence of death, reinfarction, or heart failure in stable patients with infarct-related arterial occlusion occurring 3-28 days after myocardial infarction. Furthermore, the trial suggested a tendency toward increased reinfarction with PCI during the 4-year follow-up^6^. In the DECOP trial encompassing 212 patients with initial Q-wave myocardial infarction and infarct vessel occlusion, individuals were randomly assigned to either percutaneous revascularization therapy (PTCA, n=109) or pharmacological therapy (n=103). PTCA procedures were conducted 2-15 days after symptom onset. The primary composite endpoint constituted cardiac death, nonfatal infarction, or ventricular tachyarrhythmia. The trial demonstrated that systemically advanced PTCA of the infarct vessel resulted in a higher left ventricular ejection fraction at 6 months without notable differences in clinical outcomes. However, it was associated with higher costs compared to pharmacologic therapy^7^.

In patients experiencing acute myocardial infarction with ST-segment elevation, timely reperfusion therapy, such as initial percutaneous coronary intervention (PCI) or thrombolysis, constitutes the optimal treatment. However, approximately one-third of eligible patients do not receive this early reperfusion therapy, often due to delayed presentation ^8,9^. Late-stage patients markedly differ from their early-stage counterparts. They tend to be older, more frequently female, exhibit a higher prevalence of diabetes, often necessitate transfer to a PCI center from another medical facility, and are less commonly treated with primary PCI^10^.

Clinical approaches to the late treatment of occluded infarct-related arteries in myocardial infarction past the time window for reperfusion remain varied and controversial. Late PCI poses potential risks, including procedure-related complications, myocardial injury resulting from distal embolization of atherothrombotic debris, and the potential loss of recruitable collateral flow in other coronary regions^11,12^. However, there are mechanisms through which late PCI of occluded infarct-related arteries could potentially improve outcomes. These mechanisms include the reduction of adverse left ventricular remodeling and preservation of left ventricular function^13,14^, reduction of infarct size^15^, enhancement of electrophysiologic stability, and facilitation of collateral blood vessel formation in other coronary artery regions, potentially preventing future cardiovascular events^16^.

Busk et al.^17^conducted a study comparing final infarct size (FIS) and myocardial salvage in patients undergoing primary angioplasty, distinguishing between early attendees (within 12 hours) and late attendees (between 12-72 hours) after acute myocardial infarction (AMI). They employed acute myocardial perfusion imaging (MPI) before angioplasty to evaluate the area at risk (AAR). Follow-up MPI was repeated after 30 days to assess FIS (as a percentage of left ventricular (LV) myocardium), salvage index (percentage of non-infarcted AAR), and LVEF. The findings indicated that patients with a delayed presentation (>12 hours) had a larger FIS post primary angioplasty for STEMI compared to those with an early presentation. Nevertheless, substantial myocardial salvage was observed even when the infarct-related artery was completely occluded beyond the 12-hour limit. Silva et al.^13^investigated the impact of revascularization on left ventricular (LV) remodeling and systolic function after myocardial infarction (MI) and its correlation with conservative treatment. They randomized 36 patients presenting with infarct-related artery occlusion between 12 hours and 14 days after anterior wall myocardial infarction to either undergo percutaneous coronary intervention (PCI group) or receive conservative treatment (non-PCI group). Magnetic resonance imaging was conducted at enrollment and six months later. Results indicated that late recanalization led to improvements in LVEF and myocardial contractility during late follow-up. Sustained patency of the infarct-related artery showed potential benefits in enhancing overall and local left ventricular contractility, possibly contributing to long-term prognosis post-MI. Moreover, Yousef et al.^18^demonstrated in their study that recanalization of an infarct-related artery occlusion in asymptomatic patients, approximately one month after acute myocardial infarction, tended to enhance exercise tolerance and improve the quality of life. These studies collectively suggest that late recanalization in selected patients following acute myocardial infarction could potentially contribute to salvaging myocardium, improving left ventricular function, and enhancing long-term prognosis, thereby offering potential benefits in terms of functional outcomes and quality of life.

The optimal treatment approach for patients with persistent complete occlusion of the infarct-related artery detected after the recognized reperfusion phase of myocardial infarction remains uncertain. Observational studies have consistently indicated that maintaining patency of the infarct-related artery late after myocardial infarction is linked to a reduced event rate compared to persistent occlusion^6^. In real-world scenarios, some patients experiencing ST-elevation myocardial infarction (STEMI) undergo PCI procedures beyond 48 hours after symptom onset. In this particular study, we aimed to evaluate the influence of the timing of late revascularization procedures on the long-term prognosis of patients. Notably, patients undergoing late PCI exhibited a higher thrombus load and a greater percentage necessitating intraoperative thrombus aspiration and the use of tilofiban compared to those undergoing more advanced PCI treatment. Although statistical significance was not attained, patients treated with late PCI demonstrated a trend towards a heightened risk of all-cause mortality and composite events during follow-up. Larger sample sizes in future follow-ups may provide clarity and accentuate any existing differences in outcomes between the two treatment groups.

This study has several limitations that warrant consideration: (1) Retrospective Non-randomized Design: The study’s retrospective, non-randomized nature introduces inherent limitations. Despite employing multifactorial correction for certain confounding variables, complete elimination of bias remains unattainable. Nonetheless, it’s crucial to acknowledge that this study represents a real-world scenario, providing insights into the actual clinical landscape. (2) Small Sample Size and Uncertainty: The study’s limited sample size contributes to insufficient certainty in the results. A small sample size might hinder the generalizability and robustness of findings, potentially limiting the study’s ability to draw definitive conclusions.

In summary, the late PCI group exhibited a higher thrombus load and a greater need for intraoperative tirofiban administration. However, statistical analysis revealed no significant disparity between the two groups concerning total hospitalization costs and length of hospital stay. During a follow-up period of 20 months (with a median duration of 20 months and an interquartile range of 15 to 28 months), the prognosis of patients experiencing acute ST-segment elevation myocardial infarction and undergoing PCI at either a late stage or a more advanced stage appeared comparable.

## Data Availability

Comprehensive aggregate data will be shared from the corresponding author upon request.

## Notes

### Competing Interest Statement

The authors have declared no competing interest.

### Clinical Trial

The study procedures were approved by the Medical Ethics Committee of Tongji Hospital, Tongji Medical College, Huazhong University of Science and Technology(TJ-IRB202401016).

### Funding Statement

No external funding was received.

## Reference

1. Vogel B, Claessen BE, Arnold SV, Chan D, Cohen DJ, Giannitsis E, Gibson CM, Goto S, Katus HA, Kerneis M, Kimura T, Kunadian V, Pinto DS, Shiomi H, Spertus JA, Steg PG, Mehran R. ST-segment elevation myocardial infarction. Nat Rev Dis Primers. 2019;5:39.

2. Grines CL, Mehta S. ST-segment elevation myocardial infarction management: great strides but still room for improvement. Eur Heart J. 2021;42:4550–4552.

3. Lawton JS, Tamis-Holland JE, Bangalore S, Bates ER, Beckie TM, Bischoff JM, Bittl JA, Cohen MG, DiMaio JM, Don CW, Fremes SE, Gaudino MF, Goldberger ZD, Grant MC, Jaswal JB, Kurlansky PA, Mehran R, Metkus TS, Nnacheta LC, Rao SV, Sellke FW, Sharma G, Yong CM, Zwischenberger BA. 2021 ACC/AHA/SCAI Guideline for Coronary Artery Revascularization: A Report of the American College of Cardiology/American Heart Association Joint Committee on Clinical Practice Guidelines. Circulation. 2022;145:e18–e114.

4. Thygesen K, Alpert JS, Jaffe AS, Chaitman BR, Bax JJ, Morrow DA, White HD, ESC Scientific Document Group. Fourth universal definition of myocardial infarction (2018). Eur Heart J. 2019;40:237–269.

5. Neumann F-J, Sousa-Uva M, Ahlsson A, Alfonso F, Banning AP, Benedetto U, Byrne RA, Collet J-P, Falk V, Head SJ, Jüni P, Kastrati A, Koller A, Kristensen SD, Niebauer J, Richter DJ, Seferovic PM, Sibbing D, Stefanini GG, Windecker S, Yadav R, Zembala MO, ESC Scientific Document Group. 2018 ESC/EACTS Guidelines on myocardial revascularization. Eur Heart J. 2019;40:87–165.

6. Hochman JS, Lamas GA, Buller CE, Dzavik V, Reynolds HR, Abramsky SJ, Forman S, Ruzyllo W, Maggioni AP, White H, Sadowski Z, Carvalho AC, Rankin JM, Renkin JP, Steg PG, Mascette AM, Sopko G, Pfisterer ME, Leor J, Fridrich V, Mark DB, Knatterud GL, Occluded Artery Trial Investigators. Coronary intervention for persistent occlusion after myocardial infarction. N Engl J Med. 2006;355:2395–2407.

7. Steg PG, Thuaire C, Himbert D, Carrié D, Champagne S, Coisne D, Khalifé K, Cazaux P, Logeart D, Slama M, Spaulding C, Cohen A, Tirouvanziam A, Montély J-M, Rodriguez R-M, Garbarz E, Wijns W, Durand-Zaleski I, Porcher R, Brucker L, Chevret S, Chastang C, DECOPI Investigators. DECOPI (DEsobstruction COronaire en Post-Infarctus): a randomized multi-centre trial of occluded artery angioplasty after acute myocardial infarction. Eur Heart J. 2004;25:2187–2194.

8. Eagle KA, Goodman SG, Avezum Á, Budaj A, Sullivan CM, López-Sendón J. Practice variation and missed opportunities for reperfusion in ST-segment-elevation myocardial infarction: findings from the Global Registry of Acute Coronary Events (GRACE). The Lancet. 2002;359:373–377.

9. Cohen M, Gensini GF, Maritz F, Gurfinkel EP, Huber K, Timerman A, Krzeminska-Pakula M, Santopinto J, Hecquet C, Vittori L. Prospective Evaluation of Clinical Outcomes After Acute ST-Elevation Myocardial Infarction in Patients Who Are Ineligible for Reperfusion Therapy: Preliminary Results From the TETAMI Registry and Randomized Trial. Circulation. 2003;108. doi:10.1161/01.CIR.0000091832.74006.1C.

10. Dauerman HL, Ibanez B. The Edge of Time in Acute Myocardial Infarction. J Am Coll Cardiol. 2021;77:1871–1874.

11. Verevkin A, Aspern K von, Leontyev S, Lehmann S, Borger MA, Davierwala PM. Early and Long-Term Outcomes in Patients Undergoing Cardiac Surgery Following Iatrogenic Injuries During Percutaneous Coronary Intervention. Journal of the American Heart Association: Cardiovascular and Cerebrovascular Disease. 2019;8. doi:10.1161/JAHA.118.010940.

12. G H, P K, D B, B L, M H, R S, R E. Coronary microembolization: from bedside to bench and back to bedside. Circulation. 2009;120. doi:10.1161/CIRCULATIONAHA.109.888784.

13. Silva JC, Rochitte CE, Júnior JS, Tsutsui J, Andrade J, Martinez EE, Moffa PJ, Menegheti JC, Kalil-Filho R, Ramires JF, Nicolau JC. Late coronary artery recanalization effects on left ventricular remodelling and contractility by magnetic resonance imaging. Eur Heart J. 2005;26:36–43.

14. Horie H, Takahashi M, Minai K, Izumi M, Takaoka A, Nozawa M, Yokohama H, Fujita T, Sakamoto T, Kito O, Okamura H, Kinoshita M. Long-term beneficial effect of late reperfusion for acute anterior myocardial infarction with percutaneous transluminal coronary angioplasty. Circulation. 1998;98:2377–2382.

15. Schömig A, Mehilli J, Antoniucci D, Ndrepepa G, Markwardt C, Di Pede F, Nekolla SG, Schlotterbeck K, Schühlen H, Pache J, Seyfarth M, Martinoff S, Benzer W, Schmitt C, Dirschinger J, Schwaiger M, Kastrati A, Beyond 12 hours Reperfusion AlternatiVe Evaluation (BRAVE-2) Trial Investigators. Mechanical reperfusion in patients with acute myocardial infarction presenting more than 12 hours from symptom onset: a randomized controlled trial. JAMA. 2005;293:2865–2872.

16. Monteiro P, Antunes A, Gonçalves LM, Providência LA. Long-term clinical impact of coronary-collateral vessels after acute myocardial infarction. Rev Port Cardiol. 2003;22:1051–1061.

17. Busk M, Kaltoft A, Nielsen SS, Bøttcher M, Rehling M, Thuesen L, Bøtker HE, Lassen JF, Christiansen EH, Krusell LR, Andersen HR, Nielsen TT, Kristensen SD. Infarct size and myocardial salvage after primary angioplasty in patients presenting with symptoms for <12 h vs. 12-72 h. Eur Heart J. 2009;30:1322–1330.

18. Yousef ZR, Redwood SR, Bucknall CA, Sulke AN, Marber MS. Late intervention after anterior myocardial infarction: effects on left ventricular size, function, quality of life, and exercise tolerance: results of the Open Artery Trial (TOAT Study). J Am Coll Cardiol. 2002;40:869–876.

